# Self-reported bruxism in patients with post-traumatic stress disorder

**DOI:** 10.1101/2023.09.03.23294937

**Authors:** Ana Cristina de Oliveira Solis, Felipe Corchs, Érica Panzani Duran, Cláudio Silva, Natalia Del Real, Álvaro Cabral Araújo, Yuan-Pang Wang, Francisco Lotufo-Neto

**Affiliations:** Departamento e Instituto de Psiquiatria, Hospital das Clínicas HCFMUSP, Faculdade de Medicina, Universidade de Sao Paulo, Sao Paulo, SP, BR; Departamento e Instituto de Psiquiatria (LIM-23), Hospital das Clínicas HCFMUSP, Faculdade de Medicina, Universidade de Sao Paulo, Sao Paulo, SP, BR

**Keywords:** post-traumatic stress disorder, stress, trauma, bruxism, facial pain, temporomandibular joint disorders

## Abstract

**Objective:** The present study aimed to investigate the association between self-reported awake/sleep bruxism, and orofacial pain with post-traumatic stress disorder (PTSD).

**Methods:** Participants were recruited from a university-based Trauma Ambulatory. The diagnosis of PTSD was established through a clinical interview and the Structured Clinical Interview (SCID-I/P). Thirty-eight PTSD patients and 38 controls completed the Research Diagnostic Criteria for Temporomandibular Disorders Axis-II to categorize awake/sleep bruxism and orofacial pain. Following this, we performed a short clinical examination of the temporomandibular joint and extraoral muscles.

**Results:** Adjusted logistic regression analysis showed that awake bruxism was associated with PTSD (OR = 3.38, 95% CI = 1.01-11.27, *p* = 0.047). Sleep bruxism was not associated with any covariate included in the model. In a Poisson regression model, PTSD (IRR = 3.01, 95% CI = 1.38-6.55, *p* = 0.005) and the muscle pain/ discomfort (IRR = 5.12, 95% CI = 2.80-9.36, *p* < 0.001) were significant predictors for current orofacial pain.

**Conclusions:** PTSD was associated with self-reported awake bruxism and low-intensity orofacial pain. These conditions were frequent outcomes in patients previously exposed to traumatic events.

## Introduction

Post-traumatic stress disorder (PTSD) is characterized by negative emotions and avoidance behaviors triggered after exposure to a traumatic event [1]. Data from the World Mental Health (WMH) surveys estimated that the conditional risk for PTSD after trauma exposure is approximately 4%, considering all trauma types [2]. In urban environments, the estimates are higher compared to war environments (0.1 % to 19% vs 0.8% to 5.4%). For example, events such as rape (19%), kidnapping (11%), physical abuse by a romantic partner (11.7%), or physical abuse in childhood (5.0%) are more strongly associated with PTSD than war combat experience (3.6%) [2]. However, once detected, the symptoms of PTSD last for approximately 6 years across all trauma types, especially after war combat (∼13 years) and physical abuse in childhood (∼11 years). These estimates highlight the chronic and persistent characteristics of stress in PTSD.

Bruxism is described as a repetitive activity of the masticatory system and is characterized by clenching or grinding of the teeth and/or by bracing or thrusting of the mandible. The etiology of bruxism is not completely explained [3, 4]. Current literature suggests that bruxism encompasses a broad spectrum of jaw muscle activities, centrally regulated, not peripherally, as previously supposed [5–8]. This means that overvaluation was given to morphological factors such as dental occlusion and orofacial anatomy in early theories [8]. Currently, numerous studies have confirmed that stress, anxiety, depression, sleep disturbance, medications, drugs, and certain diseases may account for this condition in different degrees, contexts, and situations. A Finnish twin cohort study also demonstrated that gene-environment interactions may explain self-reported bruxism [9].

Clinical signs of bruxism may include tooth wear, muscle pain/hypertrophy, fractured restorations, tongue indentation, “line alba” on the cheek mucosa, dentin hypersensitivity, teeth grinding sounds during sleep (usually reported by a room partner or sibling) [10], fracture of dental implants, temporomandibular disorder, orofacial pain [11], and headache. Interestingly, bruxism does not always cause a significant destructive effect on the masticatory system, such as detectable tooth wear or muscle pain [12].

The current classification system for bruxism recognizes two circadian manifestations: awake bruxism and sleep bruxism [3]. Awake bruxism (AB) is defined as “repetitive or sustained tooth contact during the daytime and/or by bracing or thrusting of the mandible and is not a movement disorder in healthy individuals” [4].

AB is frequently associated with anxiety and stress situations and involves involuntary teeth clenching or non-functional tooth contacts. This means that tooth contact is perceived during activities that require concentration, such as working or reading a book [13–16].

On the other hand, sleep bruxism (SB) is defined as “masticatory muscle activity during sleep that is characterized as rhythmic (phasic) or non-rhythmic (tonic) and is not a movement disorder or a sleep disorder in otherwise healthy individuals” [4]. SB is usually associated with alterations in the central nervous system and is correlated with micro-arousals, and it produces typical grindings sounds [17]. Clinically, AB is characterized more by clenching, while SB is more associated with both clenching and grinding [17]. Moreover, AB/SB can be detected or intensified after the use of certain psychotropic medications [4, 18].

The prevalence rates of bruxism vary according to the approach used to diagnose [4]. In adults, estimates range from 22% - 31% for AB, 12.8% for “frequent” SB, and approximately 5.5% for SB, confirmed with polysomnography [19, 20]. In general, bruxism in adults is inversely associated with age and independent of gender [15, 17]. The prevalence is higher in children. Parent-reported SB in 7-to 12-year-old children ranged from 19.5% to 36.5% [21], and in orphans, estimates were 16.9% for SB and 37.7% for clenching during daytime. [22]

In our previous study [23], we found that PTSD was associated with bruxism, but we did not differentiate between AB and SB. Considering the strength of the association between stress and bruxism, we conducted a new analysis using data from the Research Diagnostic Criteria for Temporomandibular Disorders Axis II (RDC/TMD Axis II) to clarify the effects of PTSD on self-reported AB/SB and orofacial pain. We hypothesized that patients with PTSD would have a higher frequency of AB and SB compared to control subjects. Therefore, the aim of the present study was to investigate the association between bruxism (AB ∕ SB) and orofacial pain and PTSD.

## Method

The STROBE guidelines for observational studies were followed [24].

### Study design and setting

A case-control study with a convenience sample was designed. Between October 2014 and December 2015, patients under treatment for PTSD and patients recently admitted to ambulatory studies at the Institute of Psychiatry (Ambulatory of Trauma) were invited by the psychiatrist or psychologist to participate in the dental study [25, 26]. Hospital staff and medical students without PTSD were selected for the control group. Patients and the control group were from the community-dwellers in the São Paulo metropolitan area and were contacted through advertisements in the local media or personal invitations. A total of 84 subjects were invited to participate. The final sample consisted of 76 participants. Sample size calculation and study flowchart were also described in a previous publication[23].

### Inclusion criteria

Patients should meet the PTSD criteria according to DSM-IV-TR [27]. We matched each case to one control by gender and marital status.

### Exclusion criteria

We excluded participants with [23, 25, 26]:

a. Traumatization
b. Lack of remembrance of the traumatic event
c. Abuse or dependence on alcohol or drugs
d. Subjects who exhibit any systemic disease that might hinder periodontal clinical examination.

### Ethics

The study was approved by the Ethical Committee and Research of the Medical School [23]. Written informed consent was obtained from all participants before entering the study.

### Data sources and measurements

We face-to-face interviewed through the Structured Clinical Interview (SCID-I/P). Psychologists applied the Davidson Trauma Scale [25, 26]. All patients underwent a comprehensive periodontal clinical examination and a short clinical examination of the temporomandibular joint (TMJ) and extraoral muscles by one dentist[23]. Before the dental examination, the subjects completed a general and dental health questionnaire, and the Research Diagnostic Criteria for Temporomandibular Disorders Axis II (RDC/TMD Axis II) [28]

### Tools

#### Structured Clinical Interview for DSM-IV-TR Axis I Disorder (SCID-I)

The Structured Clinical Interview for DSM-IV-TR Axis-I Disorders (SCID-I) is a standardized, semi-structured interview that is widely recognized as the gold standard for diagnosing psychiatric disorders listed in DSM-IV-TR[27]. After screening for the presence of core psychopathology, the interviewer should examine whether respondents meet criteria for affective disorders (Sections A, D), psychotic disorders (C, D), substance use disorders (SUD) (E), anxiety disorders (F), somatoform disorders (G), eating disorders (H), and adjustment disorders (I). The diagnostic strategy of the SCID-I is top-down, that is, the presence of symptoms should be screened for the supposed psychiatric diagnosis. We used the patient version of the SCID-I, which allows recording of psychiatric disorders for both current and lifetime periods. For the purposes of the present study, we only used the PTSD module (Section F). Before completing the interview, the interviewer also assessed the patients’ global assessment of functioning (GAF), that is, their best functioning in the past year. Two trained psychiatrists with experience in the treatment of anxiety disorders conducted face-to-face interviews with eligible participants. Considerable evidence of reliability and validity was found in several patients.

#### The Davidson Trauma Scale (DTS)

The DTS is a self-rating scale designed to measure symptoms of PTSD described in the DSM-IV. Respondents are inquired to rate a traumatic event according to the level of distress experienced in the last week. Particularly, DTS is composed of frequency and severity scores, each one ranging from 0 to 68 points. It is a Likert-scale and a total of 17 items are scored. The score ranges from 0 to136 [29]. Psychologists applied the Davidson Trauma Scale [26]. The DTS is not validated for the Brazilian population.

### Dental Health Questionnaire

The questionnaire consisted of self-reported questions with dichotomous responses, and was used to assess oral and systemic health, including self-rated gingival inflammation, gingival pain, bruxism, tooth mobility, use of dental floss, and mouthwashes. Open questions were designed to evaluate the frequency of dental attendance, and the use of medications. Multiple-choice questions were included to identify common systemic diseases such as diabetes, cardiovascular disease, renal disease, hepatitis, acquired immunodeficiency syndrome (AIDS).

#### Research Diagnostic Criteria for Temporomandibular Disorders Axis II (RDC/TMD Axis II)

The RDC/TMD Axis II was designed to screen patients based on depression and somatization symptoms in order to classify them according to a “chronic pain scale”. This instrument combines items from the Graded Chronic Pain Scale, Symptom Checklist −90 Revised (SCL-90-R), and limitation of the jaw [28, 30, 31].

Patients self-reported the presence of bruxism from items 15c and 15d of the RDC/TMD Axis II. *Question 15c:* “Have you been told, or do you notice that you grind your teeth or clench your jaw while sleeping at night?” and *Question 15d*: “During the day, do you grind your teeth or clench your jaw?”. These responses were dichotomous and patients should mark “*no”* or “*yes”* [28]. In our previous study [23], bruxism was detected with the following question: “Do you have the habit of grinding or clenching your teeth?”

The self-rated orofacial pain was collected from items 7, 8, and 9 of the RDC/TMD Axis II. *Question 7*: “How would you rate your facial pain on a 0 to 10 scale at the present time, that is right now, where 0 is “no pain” and 10 is “pain as bad as could be”?; *Question 8*: In the past six months, how intense was your worst pain rated on a 0 to 10 scale where 0 is “no pain” and 10 is “pain as bad as could be”?; and *Question 9:*” In the past six months, on the average, how intense was your pain rated on a 0 to 10 scale where 0 is “no pain” and 10 is “pain as bad as could be”? [That is, your usual pain at times you were experiencing pain])” [28].

#### Clinical examination of the temporomandibular joint (TMJ) and extraoral muscles

The short clinical examination included the evaluation of the opening pattern, mandibular excursive movements, TMJ sounds, and the extraoral muscle pain with digital palpation (temporalis, masseter, submandibular, and muscles of the cervical region). All examinations were performed by one trained dentist. The attending dentist was blind to psychiatric diagnosis and scores of psychometric scales.

### Variables

#### Brief explanation of nosology criteria of the international consensus on the assessment of bruxism [3, 4]

Bruxism (AB and SB) can be evaluated with non-instrumental (such as self-reported questionnaires, and clinical examination) or with instrumental approaches [such as electromyographic and audio-video recordings, polysomnography (SB), and ecological momentary assessment (EMA) app-based for AB].

A grading system was developed to determine the accuracy of the assessment, and the following levels were purposed: (1) possible awake/sleep bruxism (based on a positive self-report only), (2) probable awake/sleep bruxism (based on a positive clinical inspection, with or without a positive self-report), and (3) definite awake/sleep bruxism (based on a positive instrumental assessment, with or without a positive self-report and/or a positive clinical inspection) [4].

### Outcomes

#### Awake Bruxism (AB)

Positive self-reported for AB. Patients with AB in the present study were considered as with “possible-awake bruxism” [4].

#### Sleep Bruxism (SB)

Positive self-reported for SB. Patients with SB in the present study were considered as with “possible-sleep bruxism” [4].

#### Bruxism (B)

Positive self-reported for AB or SB or both. Patients with bruxism in the present study were considered as with “possible-sleep bruxism” or “possible-awake bruxism’ or “possible-awake ∕sleep bruxism” [4].

#### Awake and sleep bruxism (AB and SB)

Positive self-reported for AB and SB. Patients with bruxism in the present study were considered as with “possible-sleep bruxism” and with “possible-awake bruxism” [4].

#### Self-rated oral pain at the present time, in the past six months, and on average in past six months

mean scores of self-reported oral pain on a 10-item scale from RDC/TMD Axis II [28]. The mean of these three scales, multiplied by 10, characterizes the pain intensity (CPI) score [32, 33].

### Exposures considered in logistic and Poisson regression models

#### PTSD

Patients who met clinical criteria for PTSD according to DSM-IV-TR [27]

#### Muscle pain

Patients who exhibited pain or discomfort after extraoral muscle digital palpation, based on dichotomous responses (“yes” or “no”).

#### Depression

depression symptoms were evaluated with the Symptom Checklist 90-R (SCL-90-R-items) from *RDC/TMD Axis II* [30]. Scores below 0.535 were considered normal, and scores above indicated patients with moderate or severe depression [34].

### Procedures

Participants were referred to the dental service located in the same hospital to fill in the dental questionnaires and received an oral examination by one trained and calibrated examiner [23]. The psychiatric evaluation was done by two trained psychiatrists. All patients were screened by psychiatrists and members of the research group to evaluate if patients met the inclusion criteria for the psychiatric/psychological studies. A few patients received only psychotherapy. The periodontal evaluation (gingival health status) included the recording of probing pocket depth (PPD), clinical attachment level (CAL), bleeding on probing (BOP) and the frequency of plaque at six sites per tooth using a manual probe (PCPUNC, Hu-Friedy, Chicago, IL, USA). Next, participants were invited to fill out a visual analog scale (VAS) to measure the level of pain after the periodontal probing. To conclude the dental examination, subjects received a clinical examination of the temporomandibular joint (TMJ) and extraoral muscles.

### Data analysis

Firstly, we performed a descriptive analysis of socio-demographic and clinical characteristics of participants from the PTSD and control groups. The Mann-Whitney test was used to compare self-rated orofacial pain (present time, past six months, and average pain in the past six months), and the Davidson trauma scale in the PTSD and control groups. The binary logistic regression model was used to measure the association of the main dependent variables (awake or sleep bruxism, awake bruxism, awake and sleep bruxism, and sleep bruxism) with covariates (age, gender, depression, and PTSD). The Poisson or log-linear regression analysis was used to measure the effect of explanatory variables such as age, muscle pain, bruxism, and PTSD on dependent variables (present orofacial pain, past six months orofacial pain, and average orofacial pain in the past six months). Data were analyzed with STATA Statistics/Data Analysis Special Edition 15.1 (Stata Corp LLC, Texas, USA). Differences at the 5% level were considered statistically significant.

## Results

Table 1 shows the self-reported bruxism in controls and PTSD patients. The PTSD group reported a significantly higher frequency of bruxism when compared to controls (bruxism, p = 0.001; awake bruxism, p = 0.005; sleep bruxism, p = 0.002; awake and sleep bruxism, p = 0.006). Adjusted logistic regression model showed that PTSD was significantly associated with bruxism (OR = 5.34, 95% CI = 1.42-19.99, p = 0.013) and awake bruxism (OR = 3.38, 95% CI = 1.01-11.27, p = 0.047) (Table 2). Noteworthy, in these models, “awake and sleep bruxism” or “sleep bruxism” were not associated with any covariates (age, gender, depression, and PTSD) (Table 2). Table 3 depicted the self-reported orofacial pain in controls and PTSD patients. Patients with PTSD presented higher levels of orofacial pain at two different time points compared to controls (present time, p<0.001, and past six months, p<0.001, respectively), and significantly average pain in the past six months (p<0.001). Poisson regression analysis suggested that PTSD was significantly associated with orofacial pain at present time (IRR= 3.01, 95% CI= 1.38-6.55, p=0.005), past six months (IRR=2.06, 95% CI=1.28-3.31, p=0.003), and average pain in the past six months (IRR= 2.17, 95% CI= 1.30-3.61, p=0.003) (Table 4). The mean score and standard deviation (SD) of the DTS were 86.82 (SD = 30.04) and 14.61 (SD = 23.44), p < 0.001, respectively for PTSD (n = 34) and controls (n = 36). Six patients did not complete the DTS.

**Table 1.** Self-reported bruxism in no PTSD and PTSD groups.

| Variable | Controls<br>(N=37) | PTSD<br>(N=37) | *p-value* |
| --- | --- | --- | --- |
| No bruxism <sup>a</sup> | 21 (56.76%) | 7 (18.92%) | 0.001* |
| Awake or sleep bruxism <sup>a</sup> | 16 (43.24%) | 30 (81.08%) | 0.001* |
| Awake bruxism <sup>a</sup> | 12 (32.43%) | 24 (64.86%) | 0.005* |
| Sleep bruxism <sup>a</sup> | 10 (27.03%) | 23 (60.53%) | 0.002* |
| Awake and sleep bruxism <sup>a</sup> | 6 (16.22 %) | 17 (45.95%) | 0.006* |

<sup>a</sup> Pearson Chi-squared test, \* Statistically significant

**Table 2.** Adjusted odds ratios for different bruxism behavior.

| Variable | Bruxism<br>(N= 46) | Awake<br>bruxism<br>(N=36) | Sleep<br>bruxism<br>(N=33) | Awake and sleep<br>bruxism<br>(N= 23) |
| --- | --- | --- | --- | --- |
|  | OR (95% CI) | OR (95% CI) | OR (95% CI) | OR (95% CI) |
| Age | 1.0 (0.96-1.05)<br>*p*=0.817 | 1.0 (0.96-1.04)<br>*p*=0.909 | 1.02 (0.97-1.06)<br>*p*=0.339 | 1.02 (0.97-1.07)<br>*p*=0.363 |
| Gender |  |  |  |  |
| Male | 1.00 | 1.00 | 1.00 | 1.00 |
| Female | 4.22 (1.06-16.75)<br>*p*=0.041* | 2.74 (0.70-10.64)<br>*p*=0.143 | 3.69 (0.82-16.62)<br>*p*=0.089 | 3.06 (0.55-16.97)<br>*p*=0.200 |
| Depression |  |  |  |  |
| No | 1.00 | 1.00 | 1.00 | 1.00 |
| Yes | 1.15 (0.32-4.18)<br>*p*=0.822 | 1.21 (0.36-4.12)<br>*p*=0.751 | 1.93 (0.56-6.68)<br>*p*=0.297 | 2.41 (0.61-9.55)<br>*p*=0.209 |
| PTSD |  |  |  |  |
| No | 1.00 | 1.00 | 1.00 | 1.00 |
| Yes | 5.34 (1.42-19.99)<br>*p*=0.013* | 3.38 (1.01-11.27)<br>*p*=0.047* | 2.94 (0.86-9.99)<br>*p*=0.084 | 2.55 (0.68-9.56)<br>*p*=0.163 |

\* Statistically significant

**Table 3.**
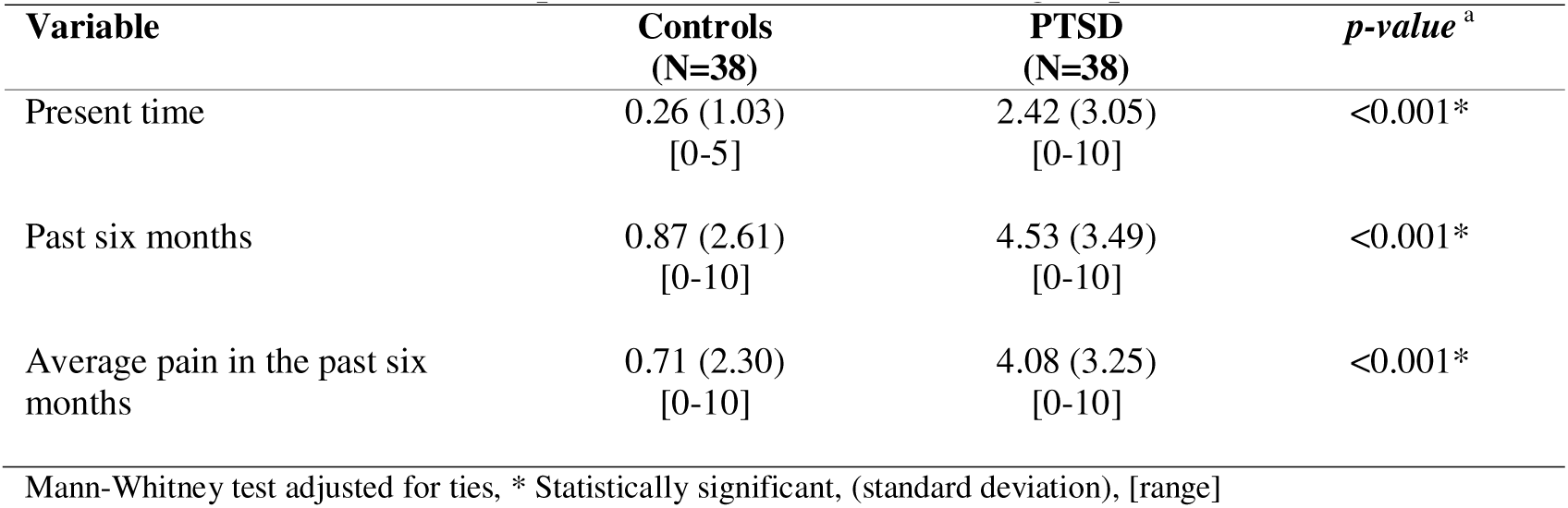
Self-rated orofacial pain in no PTSD and PTSD groups.

| Variable | Controls<br>(N=38) | PTSD<br>(N=38) | *p-value*<sup>a</sup> |
| --- | --- | --- | --- |
| Present time | 0.26 (1.03)<br>[0-5] | 2.42 (3.05)<br>[0-10] | <0.001* |
| Past six months | 0.87 (2.61)<br>[0-10] | 4.53 (3.49)<br>[0-10] | <0.001* |
| Average pain in the past six months | 0.71 (2.30)<br>[0-10] | 4.08 (3.25)<br>[0-10] | <0.001* |

Mann-Whitney test adjusted for ties, \* Statistically significant, (standard deviation), [range]

**Table 4.** Incidence rate ratios (IRRs) for different time points of orofacial pain (based on Poisson regression analysis)

| Variable | Present<br>time<br>(N=38) | Past<br>six months<br>(N=38) | Average pain in the<br>past six months<br>(N=38) |
| --- | --- | --- | --- |
|  | IRRs (95% CI) | IRRs (95% CI) | IRRs (95% CI) |
| Depression |  |  |  |
| No | 1.00 | 1.00 | 1.00 |
| Yes | 1.37 (0.70-2.67)<br>*p*=0.358 | 1.45 (0.91-2.29)<br>*p*=0.112 | 1.45 (0.89-2.35)<br>*p*=0.131 |
| Muscle pain |  |  |  |
| No | 1.00 | 1.00 | 1.00 |
| Yes | 5.12 (2.80-9.36)<br>*p*<0.001* | 1.86 (1.31-2.66)<br>*p*=0.001* | 2.22 (1.52-3.24)<br>*p*<0.001* |
| Bruxism |  |  |  |
| No | 1.00 | 1.00 | 1.00 |
| Yes | 1.17 (0.62-2.19)<br>*p*=0.625 | 2.17 (1.34-3.53)<br>*p*=0.002* | 2.28 (1.34-3.86)<br>*p*=0.002* |
| PTSD |  |  |  |
| No | 1.00 | 1.00 | 1.00 |
| Yes | 3.01 (1.38-6.55)<br>*p*=0.005* | 2.06 (1.28-3.31)<br>*p*=0.003* | 2.17 (1.30-3.61)<br>*p*=0.003* |

\* Statistically significant

## Discussion

We observed that PTSD was associated with self-reported AB in a sample composed especially of Caucasian female adults exposed to traumatic events related to urban violence. Moreover, PTSD was also associated with incidence rate ratios of present and past six-months orofacial pain. Indeed, our results agree with most studies that have evaluated the effects of stress and anxiety on bruxism and on orofacial pain [13, 14, 35–40].

Overall, the present study describes a specific activity of the masticatory system in patients previously exposed to traumatic events in the urban environment. Interestingly, despite all patients receiving psychiatry and psychotherapy treatments, the impact of such exposure at the oral level was reported by most patients. During the oral examination, particularly muscle digital palpation and periodontal clinical examination, PTSD patients were more sensitive to pain and self-reported awake bruxism. Importantly, the trauma exposures were validated by clinical experts. In the field of PTSD, such clinical inspection is quite relevant.

Our study demonstrated that PTSD was associated with the likelihood of AB (Adjusted OR=3.38, 95% CI= 1.01-11.27, *p*=0.047), but not for SB. Similar AB-results were described for exposures such as social phobia (Crude OR=23.7, 95% CI=2.94-190.6, *p*=0.003) [35], anxiety symptoms (Adjusted OR=2.2, 95% CI=1.24-3.9, *p*=0.007) [13], and sexual assault (Adjusted OR=2.26, 95% CI=1.44-3.56, *p*<0.05)[41]. In a Finnish employee population, bruxism was significantly associated with severe stress experience (OR=5.00, 95% CI=2.84-8.82, *p*<0.001) [42]. Awake bruxism is a repetitive masticatory muscle activity initially developed unconsciously [43]. This activity is characterized by non-functional tooth contacts, teeth clenching (a marked tooth contact with tense jaw muscles), teeth grinding, and jaw clenching without teeth contact [15]. Curiously, some authors have reported that even long-lasting and low-intensity tooth-clenching may also be associated with masticatory muscle pain [44, 45]. Studies using ecological momentary assessment, i.e., evaluation of oral behaviors in the natural environment with questions sent by email, microprocessor-controlled recorder, or pagers during the day, observed that the frequency of non-functional tooth contacts was higher in patients with muscle pain and temporomandibular disorder compared to controls [46, 47]. Therefore, AB can trigger long-lasting muscle contraction or high-intensity muscle contractions that may injure muscle fibers [48, 49]. In addition, the stress experience can induce hyperalgesia and modify the pain threshold through numerous neurobiological changes [50].

Interesting, in the present study, PTSD was not a significant exposure for SB in multivariable analysis (Adjusted OR=2.75, 95% CI= 0.82-9.26, *p*=1.101). In a recent Dutch study conducted in 673 patients diagnosed with severe PTSD, only physical assault was found to be associated with SB (Adjusted OR=2.06, 95% CI=1.04-4.08, *p*<0.05) among 19 types of traumatic events experienced [41]. Most epidemiological data on bruxism is based on self-reports, and the assessment of self-reported bruxism with psychological variables was encouraged [4]. Nevertheless, the definitive criteria for SB incorporate the utilization of polysomnography with audio/video recordings and clinical examination [3, 4]. Our findings should be interpreted considering this possible bias. Secondly, our sample was composed of highly medicated PTSD patients. This means that some patients also received pharmacological treatment for sleep disturbance. Despite these clinical considerations, our non-significant SB-result is comparable to those described for anxiety symptoms (Adjusted OR=1.53, 95% CI=0.89-2.61, *p*=0.11), social phobia, and self-reported stress [13, 35, 36, 39]. SB is characterized by typical rhythmic masticatory muscle activity correlated with sleep arousals; and appears to be more associated in the adult population with autonomic sympathetic cardiac activity, gastroesophageal reflux disease, a history of SB in childhood than psychological stress per se [51].

Orofacial pain is related to dysfunction affecting motor and sensory transmission in the trigeminal nerve system. Many studies have reported higher frequencies of orofacial pain in patients with PTSD [40, 52]. In the present study, PTSD was a significant predictor for present and past six-months orofacial pain, considering depression symptoms, muscle pain (detected clinically), and self-reported bruxism as covariates. Our results agree with studies conducted in German and Dutch population where PTSD was a significant predictor for joint and muscle pain [40, 41].

Altered pain has been associated with PTSD, sometimes in opposite directions. Increased pain sensitivity was associated with anxiety symptoms and pain catastrophizing, and increased pain threshold with dissociative symptoms. Whereas defensive reactions to threats involve an overall increase in threat sensitivity [53], which may explain increased clinical pain in disorders such as PTSD [53], some phases of the defensive spectrum require analgesia and endogenous opioid release to focus resources on the external threat [54]. Sensitivity to pain may adaptively increase in post-attack safer situations, in which attention can be given to the wounds, or when threat potential is sustained and unpredictable, such that pain can hinder risk-taking foraging. Furthermore, the long-term PTSD was associated with a higher catastrophic and frightful orientation to bodily signals [55]. These findings can explain why patients with PTSD also exhibited higher scores of self-reported “orofacial pain in the past six months” and the “average pain in the past six months” (Table 4).

One important strength of the present study was the evaluation of PTSD by clinical psychiatrists compared to self-reported evaluation. Still, the study design also contributed to exploring the significant exposures associated with bruxism activities and orofacial pain. On the other hand, the effect of medication was not controlled in the statistical analysis. Additionally, for a complementary assessment of AB/SB, there are recommendations of an instrumental approach, such as using smartphone alerts during the daytime to detect AB in the natural environment [15], and polysomnography with audio/video recordings for patients with sleep bruxism. Nevertheless, the odds of our data were similar to those reported in population-based and clinical studies.

Finally, patients exposed to traumatic events related to urban violence and who develop PTSD exhibited an increased risk for self-reported awake bruxism and orofacial pain compared to controls. We suggest including a two-question screening for bruxism in psychiatry/psychology interviews to improve under-identification and to prevent harmful consequences at the orofacial level. A well-balanced psychiatry/psychology interview will determine the appropriate time for the patient to undergo a dental evaluation, if necessary. Furthermore, the estimates of PTSD among the urban population highlight the importance of the dental team considering significant and unexposed emotional factors during routine dental examinations. Patients who report high stress should be referred to a mental health professional to rule out possible interference of psychological problems in dental treatment and outcomes. Finally, more studies in other countries and in different age cohorts should be designed to better clarify this issue.

## Data Availability

All data produced in the present study are available upon reasonable request to the authors

## Author Contributions

**Concept and design:** AC Solis and F Lotufo-Neto. **Data acquisition:** AC Solis, EP Duran, C. Silva, N Del Real, AC Araújo and F Lotufo-Neto. **Analysis, or interpretation of data**: AC Solis, F Corchs, Y-P Wang and F Lotufo-Neto. AC Solis had full access to all the data in the study and takes responsibility for the integrity of the data and the accuracy of the data analysis. **Drafting of the manuscript:** AC Solis, F Corchs, Y-P Wang, and F Lotufo-Neto. **Critical revision of the manuscript for important intellectual content:** All authors. **Supervision:** F Lotufo-Neto. AC Solis conducted the study as a part of her postdoctoral research at the FMUSP. All authors approved the final version of the manuscript for submission.

## Institutional Review Board Statement

This project was approved by the Ethical Committee and Research of the Department and Institute of Psychiatry, Medical School, University of São Paulo (IPq-FMUSP), São Paulo, Brazil (CAAE:33296714.7.0000.0068).

## Informed Consent Statement

All participants received instructions about the study, read and signed the informed consent.

## References

1. Association, A.P., *Diagnostic and statistical manual of mental disorders*. Fifth edition. ed. Diagnostic and statistical manual of mental disorders : DSM-5. 2013, Arlington, VA: American Psychiatric Association.

2. Kessler, R.C., et al., Trauma and PTSD in the WHO World Mental Health Surveys. Eur J Psychotraumatol, 2017. 8(sup5): p. 1353383.

3. Lobbezoo, F., et al., Bruxism defined and graded: an international consensus. J Oral Rehabil, 2013. 40(1): p. 2–4.

4. Lobbezoo, F., et al., International consensus on the assessment of bruxism: Report of a work in progress. J Oral Rehabil, 2018.

5. Manfredini, D., et al., The bruxism construct: From cut-off points to a continuum spectrum. J Oral Rehabil, 2019. 46(11): p. 991–997.

6. Lobbezoo, F. and M. Naeije, Bruxism is mainly regulated centrally, not peripherally. J Oral Rehabil, 2001. 28(12): p. 1085–91.

7. Behr, M., et al., The two main theories on dental bruxism. Ann Anat, 2012. 194(2): p. 216–9.

8. Manfredini, D., et al., Occlusal factors are not related to self-reported bruxism. J Orofac Pain, 2012. 26(3): p. 163–7.

9. Ahlberg, J., et al., Correlates and genetics of self-reported sleep and awake bruxism in a nationwide twin cohort. J Oral Rehabil, 2020. 47(9): p. 1110–1119.

10. Macedo, C.R., et al., Pharmacotherapy for sleep bruxism. Cochrane Database Syst Rev, 2014(10): p. CD005578.

11. van Selms, M.K., et al., Are Pain-Related Temporomandibular Disorders the Product of an Interaction Between Psychological Factors and Self-Reported Bruxism? J Oral Facial Pain Headache, 2017. 31(4): p. 331–338.

12. Pergamalian, A., et al., The association between wear facets, bruxism, and severity of facial pain in patients with temporomandibular disorders. J Prosthet Dent, 2003. 90(2): p. 194–200.

13. Tavares, L.M., et al., Cross-sectional study of anxiety symptoms and self-report of awake and sleep bruxism in female TMD patients. Cranio, 2016. 34(6): p. 378–381.

14. Ahlberg, J., et al., Self-reported bruxism mirrors anxiety and stress in adults. Med Oral Patol Oral Cir Bucal, 2013. 18(1): p. e7–11.

15. Bracci, A., et al., Frequency of awake bruxism behaviours in the natural environment. A 7-day, multiple-point observation of real-time report in healthy young adults. J Oral Rehabil, 2018. 45(6): p. 423–429.

16. Paesani, D., *Bruxism: Theory and Practice*. 2010, United Kingdom: Quintessence Publishing Co. Ltd. 540.

17. Khoury, S., et al., Sleep Bruxism-Tooth Grinding Prevalence, Characteristics and Familial Aggregation: A Large Cross-Sectional Survey and Polysomnographic Validation. Sleep, 2016. 39(11): p. 2049–2056.

18. Melo, G., et al., Association between psychotropic medications and presence of sleep bruxism: A systematic review. J Oral Rehabil, 2018. 45(7): p. 545–554.

19. Manfredini, D., et al., Epidemiology of bruxism in adults: a systematic review of the literature. J Orofac Pain, 2013. 27(2): p. 99–110.

20. Maluly, M., et al., Polysomnographic study of the prevalence of sleep bruxism in a population sample. J Dent Res, 2013. 92(7 Suppl): p. 97S–103S.

21. van Selms, M.K.A., et al., Geographical variation of parental-reported sleep bruxism among children: comparison between the Netherlands, Armenia and Indonesia. Int Dent J, 2018.

22. Friedman Rubin, P., et al., Prevalence of bruxism and temporomandibular disorders among orphans in southeast Uganda: A gender and age comparison. Cranio, 2018. 36(4): p. 243–249.

23. de Oliveira Solis, A.C., et al., Impact of post-traumatic stress disorder on oral health. J Affect Disord, 2017. 219: p. 126–132.

24. von Elm, E., et al., The Strengthening the Reporting of Observational Studies in Epidemiology (STROBE) statement: guidelines for reporting observational studies. J Clin Epidemiol, 2008. 61(4): p. 344–9.

25. Araujo, A.C., et al., Traumatic memory retrieval followed by electroconvulsive therapy as a treatment for posttraumatic stress disorder: A pilot study. Psychiatry Res, 2023. 326: p. 115353.

26. Duran, E.P., et al., A randomized clinical trial to assess the efficacy of trial-based cognitive therapy compared to prolonged exposure for post-traumatic stress disorder: preliminary findings. CNS Spectr, 2021. 26(4): p. 427–434.

27. First, M.B., et al., *Structured Clinical Interview for DSM-IV-TR axis I disorders, research version, patient edition (SCID-I/P)*. 2002, New York: New York State Psychiatric Institute, Biometrics Research Department.

28. de Lucena, L.B., et al., Validation of the Portuguese version of the RDC/TMD Axis II questionnaire. Braz Oral Res, 2006. 20(4): p. 312–7.

29. Davidson, J.R., et al., Assessment of a new self-rating scale for post-traumatic stress disorder. Psychol Med, 1997. 27(1): p. 153–60.

30. Ohrbach, R., et al., The Research Diagnostic Criteria for Temporomandibular Disorders. IV: evaluation of psychometric properties of the Axis II measures. J Orofac Pain, 2010. 24(1): p. 48–62.

31. Schiffman, E., et al., Diagnostic Criteria for Temporomandibular Disorders (DC/TMD) for Clinical and Research Applications: recommendations of the International RDC/TMD Consortium Network* and Orofacial Pain Special Interest Group†. J Oral Facial Pain Headache, 2014. 28(1): p. 6–27.

32. Von Korff, M., et al., Grading the severity of chronic pain. Pain, 1992. 50(2): p. 133–49.

33. Dworkin, S.F., et al., Measurement of characteristic pain intensity in field research. Pain, 1990. 41: p. S290.

34. Manfredini, D., et al., Axis II psychosocial findings predict effectiveness of TMJ hyaluronic acid injections. Int J Oral Maxillofac Surg, 2013. 42(3): p. 364–8.

35. Hermesh, H., et al., Bruxism and oral parafunctional hyperactivity in social phobia outpatients. J Oral Rehabil, 2015. 42(2): p. 90–7.

36. Manfredini, D., et al., Assessment of Anxiety and Coping Features in Bruxers: A Portable Electromyographic and Electrocardiographic Study. J Oral Facial Pain Headache, 2016. 30(3): p. 249–54.

37. Sherman, J.J., et al., Post-traumatic stress disorder among patients with orofacial pain. J Orofac Pain, 2005. 19(4): p. 309–17.

38. De Leeuw, R., et al., Prevalence of post-traumatic stress disorder symptoms in orofacial pain patients. Oral Surg Oral Med Oral Pathol Oral Radiol Endod, 2005. 99(5): p. 558–68.

39. Ohlmann, B., et al., Are there associations between sleep bruxism, chronic stress, and sleep quality? J Dent, 2018. 74: p. 101–106.

40. Kindler, S., et al., Association Between Symptoms of Posttraumatic Stress Disorder and Signs of Temporomandibular Disorders in the General Population. J Oral Facial Pain Headache, 2018. 33(1): p. 67–76.

41. Knibbe, W., et al., Prevalence of painful temporomandibular disorders, awake bruxism and sleep bruxism among patients with severe post-traumatic stress disorder. J Oral Rehabil, 2022. 49(11): p. 1031–1040.

42. Ahlberg, J., et al., Reported bruxism and stress experience. Community Dent Oral Epidemiol, 2002. 30(6): p. 405–8.

43. Iida, T., P.B. Fenwick, and A.A. Ioannides, Analysis of brain activity immediately before conscious teeth clenching using magnetoencephalographic method. J Oral Rehabil, 2007. 34(7): p. 487–96.

44. Farella, M., et al., Jaw muscle soreness after tooth-clenching depends on force level. J Dent Res, 2010. 89(7): p. 717–21.

45. Cioffi, I., et al., Frequency of daytime tooth clenching episodes in individuals affected by masticatory muscle pain and pain-free controls during standardized ability tasks. Clin Oral Investig, 2017. 21(4): p. 1139–1148.

46. Funato, M., et al., Evaluation of the non-functional tooth contact in patients with temporomandibular disorders by using newly developed electronic system. J Oral Rehabil, 2014. 41(3): p. 170–6.

47. Chen, C.Y., et al., Nonfunctional tooth contact in healthy controls and patients with myogenous facial pain. J Orofac Pain, 2007. 21(3): p. 185–93.

48. Manfredini, D., et al., Current Concepts of Bruxism. Int J Prosthodont, 2017. 30(5): p. 437–438.

49. Endo, H., et al., Clenching occurring during the day is influenced by psychological factors. J Prosthodont Res, 2011. 55(3): p. 159–64.

50. Jennings, E.M., et al., Stress-induced hyperalgesia. Prog Neurobiol, 2014. 121: p. 1–18.

51. Castroflorio, T., et al., Sleep bruxism and related risk factors in adults: A systematic literature review. Arch Oral Biol, 2017. 83: p. 25–32.

52. Muhvić-Urek, M., et al., Oral health status in war veterans with post-traumatic stress disorder. J Oral Rehabil, 2007. 34(1): p. 1–8.

53. Corchs, F., et al., Sensitivity to aversive stimulation, posttraumatic symptoms and migraines: what do they have in common? Med Hypotheses, 2011. 77(4): p. 534–5.

54. Fanselow, M.S. and L.S. Lester, A functional behavioristic approach to aversively motivated behavior: Predatory imminence as a determinant of the topography of defensive behavior, in Evolution and learning. 1988, Lawrence Erlbaum Associates, Inc: Hillsdale, NJ, US. p. 185–212.

55. Tsur, N., et al., The traumatized body: Long-term PTSD and its implications for the orientation towards bodily signals. Psychiatry Res, 2018. 261: p. 281–289.

